# Prevalence of hypertension and controlled hypertension among United States adults: Evidence from NHANES 2017-18 survey

**DOI:** 10.1101/2020.07.27.20162628

**Authors:** Juwel Rana, John Oldroyd, Md. Momin Islam, Carla E Tarazona-Meza, Rakibul M. Islam

## Abstract

**Background:** This study aims to compare the prevalence of hypertension (HTN) and controlled hypertension (CHTN) in US adults and determine the absolute difference in the prevalence of HTN and CHTN between the JNC7 and ACC/AHA 2017 guidelines.

**Methods:** Data for this study were derived from the most recent cycle of the National Health and Nutrition Examination Survey (NHANES) 2017-2018. After excluding participants with missing systolic blood pressure (BP) or diastolic BP and aged <18 years, 4730 participants were included in the final analyses. BP was defined as the average of the first three measurements. The prevalence of HTN and CHTN, including absolute differences of these prevalences, were estimated using both JNC7 and ACC/AHA 2017 guidelines.

**Results:** The overall weighted prevalence of HTN was 31.7% (95% CI: 28.7, 34.8) based on JNC7, while the corresponding prevalence was 45.6% (95% CI: 43.0, 48.3) when new guideline of ACC/AHA was used. Of the people who had HTN according to the JNC7 and ACC/AHA 2017 guidelines, 48.2% (95% CI: 44.4-52.0) and 21.0% (95% CI: 18.1-24.2) had a controlled blood pressure level, respectively. When blood pressure was assessed using both guidelines, the greatest absolute increase in rates of HTN and CHTN was 17.4% and 30.0% in people aged 40-59 years, respectively.

**Conclusion:** Given the high burden of disease due to complications arising from untreated HTN, as well as the higher costs of untreated disease, new guidelines have important public health implications to early detection of patients at risk and prevent complications across different populations.

## 1. Introduction

The American College of Cardiology/American Heart Association (ACC/AHA) have developed evidence-based guidelines for the prevention and first line management of hypertension (HTN) in the US [1]. They were developed by experts and translate the best available evidence into guidelines for clinical practice. Although they are written for a US population, they have global impact. Despite this, HTN remains a significant contributor to the global burden of disease, and it remains an important risk factor for cardiovascular morbidity and mortality [2–4]. HTN has previously been defined by the Seventh Report of the Joint National Committee on Prevention, Detection, Evaluation, and Treatment of High Blood Pressure (JNC7) as individuals who have a systolic BP ≥140 mmHg or a diastolic BP ≥90 mmHg or take any prescribed drugs to control BP [5]. However, the ACC/AHA 2017 guidelines lowered the systolic and diastolic blood pressure thresholds for HTN [1]. According to the ACC/AHA 2017 guidelines, individuals who have a systolic BP ≥130 mmHg or a diastolic BP ≥80 mmHg or take any prescribed drugs to control BP were categorized as hypertensive. Application of the ACC/AHA 2017 guidelines would reclassify many individuals who had normal blood pressure (or were prehypertensive) using JNC7 criteria, as hypertensive. While this classification of people as hypertensive at lower levels of blood pressure would allow earlier intervention in the natural history of the disease, it also has implications for the evaluation of the outcomes of current prevention and control methods.

Previous studies that have investigated the impact of the new ACC/AHA 2017 guidelines have found marked increases in prevalence rates. For example, when blood pressure was assessed using both JNC7 and ACC/AHA 2017 guidelines among reproductive-aged US women, the prevalence of HTN increased by 112% [2]. This has significant implications for blood pressure management in the US. The impact of the new guidelines has also been examined in other populations in the US [6,7]. In these studies, an absolute increase in prevalence varied from 14.7% to 26.8% in the US when ACC/AHA 2017 guidelines were applied [7]. Repeating this analysis using more recent data will provide up to date evidence for planning future health policy interventions in the management of HTN.

In this study, we aim to use the most recent cycle of NHANES 2017-2018 data to compare the prevalence of HTN and controlled hypertension (CHTN) in US adults and determine the absolute difference in the prevalence of HTN and CHTN between the JNC7 and ACC/AHA 2017 guidelines.

## 2. Material and methods

### 2.1 Design, setting, and participants

Data for this study were derived from the National Health and Nutrition Examination Survey (NHANES) 2017–2018, which was conducted by the National Center for Health Statistics of the US Centers for Disease Control and Prevention. To ensure national representativeness, the study samples are identified through a complex, stratified, multistage probability sampling design. It consists of a four-stage sample: counties, segments, households, and individuals. Survey participants completed in-home interviews and then visited a mobile examination center, where they responded to additional questionnaires and underwent a medical examination and blood sample collection. NHANES maintains high standards to ensure minimal non-sampling and measurement errors during survey planning, data collection, and processing. After excluding participants with missing systolic BP or diastolic BP and aged <18 years, 4730 participants were included in the final analyses. Participants completed standardized questionnaires that assessed demographics; prior diagnosis of HTN and antihypertensive medication use; prior diagnosis of high blood cholesterol; and the use of lipid-lowering drugs; and diabetes mellitus. Body mass index was calculated as weight in kilograms divided by height in meters squared. Diabetes mellitus was self-reported, defined by a positive response to any of the questions, “Have you ever been told by a doctor that you have diabetes?”; “Are you now taking insulin?”; “Are you now taking diabetes pills to lower your blood sugar?” The study was approved by the National Center for Health Statistics Research Ethics Review Board, and all adult participants provided written informed consent [8].

### 2.2 Blood pressure measurement

During the medical examination, BP was measured after resting quietly in a seated position for 5 minutes, and after the participant’s maximum inflation level has been determined. Both mean systolic BP and mean diastolic BP were defined as the average of the first three measurements. We did not include the fourth measurement because of the large number of missing values. All BP measurements were taken in the mobile examination center using the right arm unless specific conditions prohibit the use of the right arm. Prior to BP measurements, upper arm circumference is measured, which is done to guide the selection of cuff size.

### 2.3 Definition of hypertension and control

According to the JNC7 guideline, individuals who have a systolic BP ≥140 mmHg or a diastolic BP ≥90 mmHg or take any prescribed drugs to control BP were categorized as hypertensive. According to the ACC/AHA 2017 guideline, individuals who have a systolic BP ≥130 mmHg or a diastolic BP ≥80 mmHg or take any prescribed drugs to control BP were categorized as hypertensive. According to the JNC7 guideline, CHTN is defined if individuals have systolic BP<140 mmHg and diastolic BP<90 mmHg among those with HTN. According to ACC/AHA 2017 guideline, CHTN is defined if individuals have systolic BP<130 mmHg and diastolic BP< 80 mmHg among those with HTN.

### 2.4 Statistical Analysis

The characteristics of the study participants were presented as frequencies and percentages or mean (±standard deviation; SD). The prevalence of HTN and CHTN were estimated using both JNC7 and ACC/AHA 2017 guidelines. We then estimated the absolute differences for the prevalence of HTN and CHTN between the ACC/AHA 2017 and JNC7 guidelines. All prevalences and absolute differences were reported with 95% confidence intervals (CIs). We conducted weighted analysis to adjust for the clustered sampling design of the survey. Analyses were performed using statistical software Stata version 16.0 [9], and codes are available upon request.

## 3. Results

A total of 4730 weighted participants were included in the analysis. The mean (±SD) age of the participants was 49.5 (±18.6) (Table 1). Of the study participants, 51.2% (2425) were female, 62.5% (1638) were white, 62.7% (2669) were married or living together, 42.3% (1953) were obese and 11.5% (771) had diabetes. Overall, the mean (± SD) systolic BP and diastolic BP were 125.4 (± 19.3) and 71.6 (± 12.5) mmHg, respectively. The mean (± SD) total cholesterol, triglyceride, high-density lipoprotein, and low-density lipoprotein were 187.1 (±41.5), 4.8 (±0.6), 53.2 (±15.6), and 105.3 (±37.0), respectively.

**Table 1:**
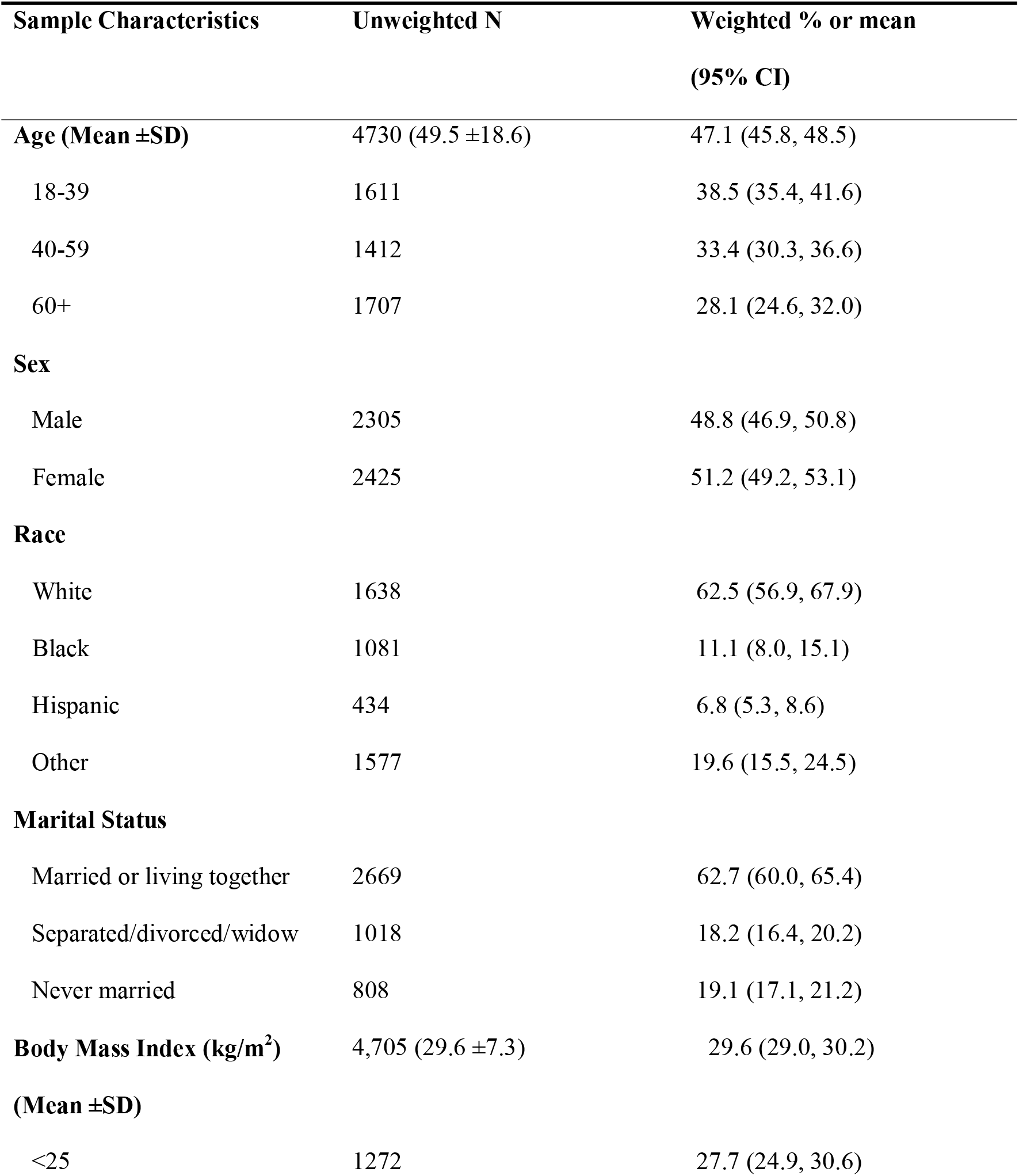
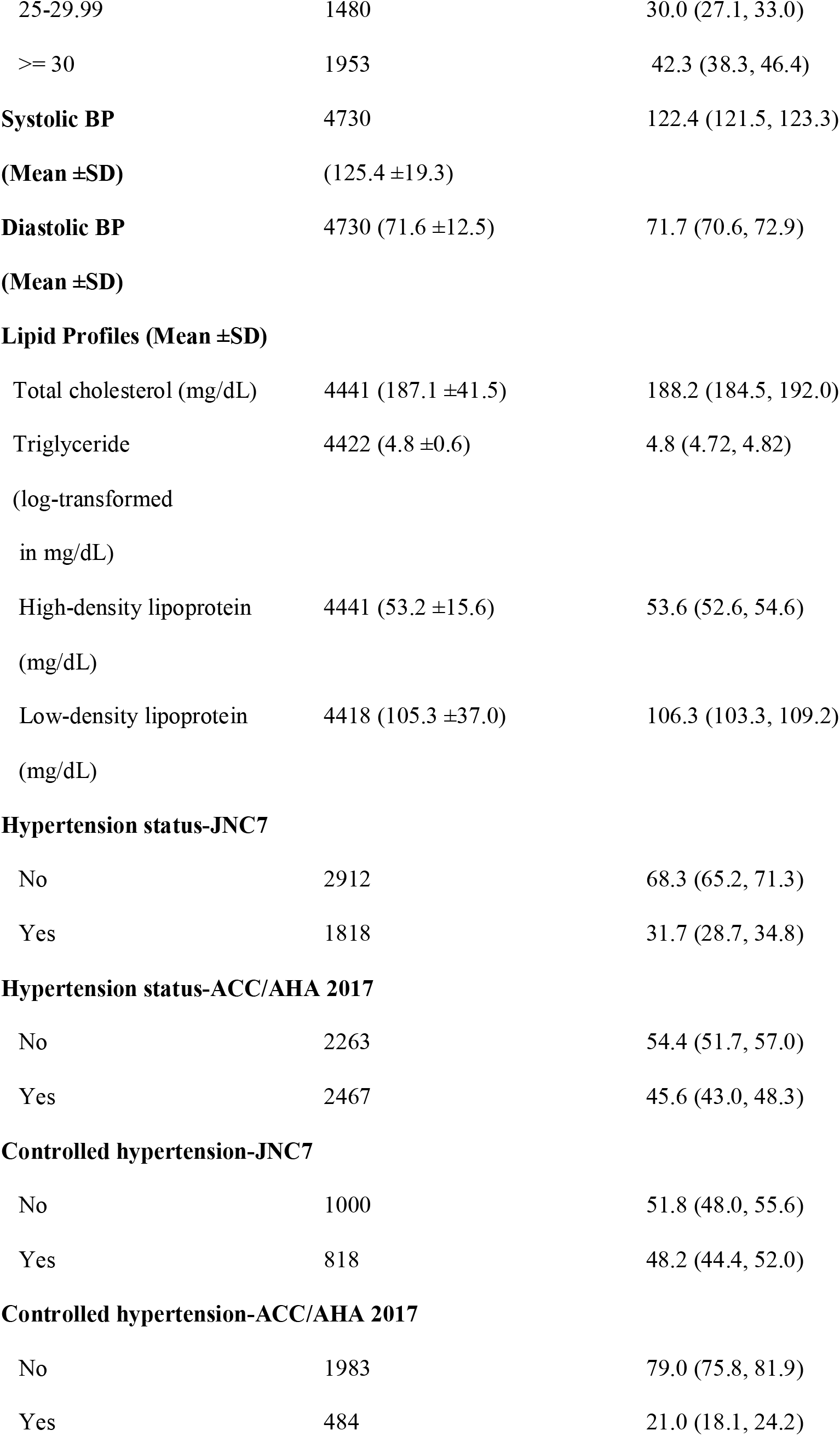
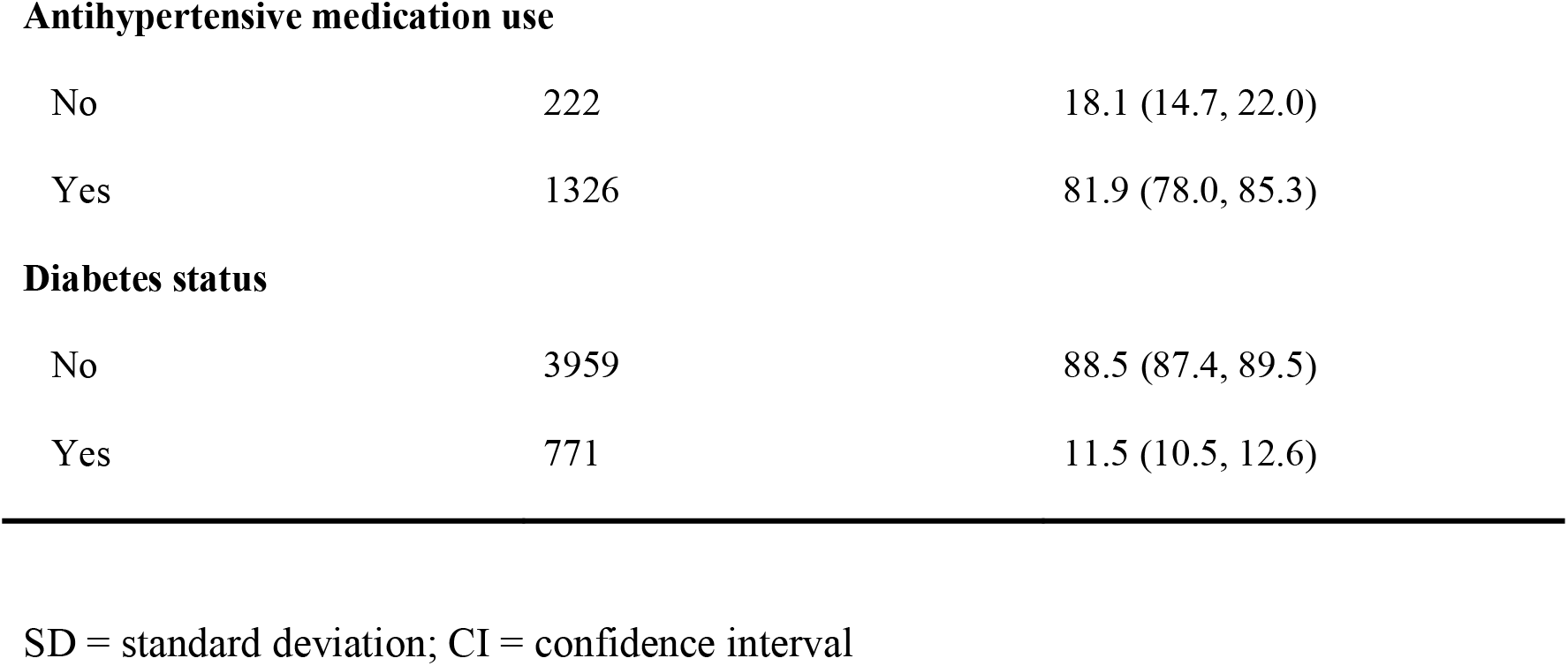
Characteristics of US adults in the NHANES 2017-18 (Unweighted Sample and weighted percentage)

| **Sample Characteristics** | **Unweighted N** | **Weighted % or mean**<br>**(95% CI)** |
| --- | --- | --- |
| **Age (Mean ±SD)** | 4730 (49.5 ±18.6) | 47.1 (45.8, 48.5) |
| 18-39 | 1611 | 38.5 (35.4, 41.6) |
| 40-59 | 1412 | 33.4 (30.3, 36.6) |
| 60+ | 1707 | 28.1 (24.6, 32.0) |
| **Sex** |  |  |
| Male | 2305 | 48.8 (46.9, 50.8) |
| Female | 2425 | 51.2 (49.2, 53.1) |
| **Race** |  |  |
| White | 1638 | 62.5 (56.9, 67.9) |
| Black | 1081 | 11.1 (8.0, 15.1) |
| Hispanic | 434 | 6.8 (5.3, 8.6) |
| Other | 1577 | 19.6 (15.5, 24.5) |
| **Marital Status** |  |  |
| Married or living together | 2669 | 62.7 (60.0, 65.4) |
| Separated/divorced/widow | 1018 | 18.2 (16.4, 20.2) |
| Never married | 808 | 19.1 (17.1, 21.2) |
| **Body Mass Index (kg/m<sup>2</sup>)** | 4,705 (29.6 ±7.3) | 29.6 (29.0, 30.2) |
| **(Mean ±SD)** |  |  |
| <25 | 1272 | 27.7 (24.9, 30.6) |
| 25-29.99 | 1480 | 30.0 (27.1, 33.0) |
| >= 30 | 1953 | 42.3 (38.3, 46.4) |
| **Systolic BP** | 4730 | 122.4 (121.5, 123.3) |
| **(Mean ±SD)** | (125.4 ±19.3) |  |
| **Diastolic BP** | 4730 (71.6 ±12.5) | 71.7 (70.6, 72.9) |
| **(Mean ±SD)** |  |  |
| **Lipid Profiles (Mean ±SD)** |  |  |
| Total cholesterol (mg/dL) | 4441 (187.1 ±41.5) | 188.2 (184.5, 192.0) |
| Triglyceride | 4422 (4.8 ±0.6) | 4.8 (4.72, 4.82) |
| (log-transformed<br>in mg/dL) |  |  |
| High-density lipoprotein | 4441 (53.2 ±15.6) | 53.6 (52.6, 54.6) |
| (mg/dL) |  |  |
| Low-density lipoprotein | 4418 (105.3 ±37.0) | 106.3 (103.3, 109.2) |
| (mg/dL) |  |  |
| **Hypertension status-JNC7** |  |  |
| No | 2912 | 68.3 (65.2, 71.3) |
| Yes | 1818 | 31.7 (28.7, 34.8) |
| **Hypertension status-ACC/AHA 2017** |  |  |
| No | 2263 | 54.4 (51.7, 57.0) |
| Yes | 2467 | 45.6 (43.0, 48.3) |
| **Controlled hypertension-JNC7** |  |  |
| No | 1000 | 51.8 (48.0, 55.6) |
| Yes | 818 | 48.2 (44.4, 52.0) |
| **Controlled hypertension-ACC/AHA 2017** |  |  |
| No | 1983 | 79.0 (75.8, 81.9) |
| Yes | 484 | 21.0 (18.1, 24.2) |
| **Antihypertensive medication use** |  |  |
| No | 222 | 18.1 (14.7, 22.0) |
| Yes | 1326 | 81.9 (78.0, 85.3) |
| **Diabetes status** |  |  |
| No | 3959 | 88.5 (87.4, 89.5) |
| Yes | 771 | 11.5 (10.5, 12.6) |

SD = standard deviation; CI = confidence interval

### 3.1 Hypertension prevalence

The overall weighted prevalence of HTN among US adults was 31.7% (95% CI: 28.7, 34.8) based on JNC7, while the corresponding prevalence was 45.6% (95% CI: 43.0, 48.3) when new guideline of ACC/AHA was used. Of the people who had HTN according to the JNC7 and ACC/AHA 2017 guidelines, 48.2% (95%CI: 44.4-52.0) and 21.0% (95%CI 18.1-24.2) had a controlled blood pressure level, respectively.

Table 2 summarized the prevalence of HTN and the absolute difference by age, sex, ethnicity, BMI, and diabetes status, comparing the previous guideline with the new guideline. According to ACC/AHA 2017 guideline, the prevalence was the highest among individuals aged 60 years and above (73.2%, 95% CI: 71.1-75.3), followed by 40-59 years (51.2%, 95% CI: 48.6-53.8), and 18-39 years (20.6%, 95% CI: 18.7-22.7). The corresponding prevalence based on JNC7 were 63.0% (61.0-65.0) for 60 years and above age group, 33.8% (31.3-36.3) for 40-59 age group and 7.0 % (5.8 −8.4) for 18-39 years age group. The highest absolute difference between the two guidelines was 17.4 % (16.7-17.5) in the middle age group. According to the 2017 ACC/AHA classification, more than half of the male respondents had HTN (51.1%, 95% CI: 49.0-53.2), compared with 40.4% (95% CI: 38.4-42.4) of the female respondents. The prevalence of HTN, according to JNC7, was 33.1% (95% CI: 31.2-35.1) among males and 30.3% (95% CI: 28.5-32.2) among females. Male (18.0%, 95% CI: 17.8-18.1) had a higher absolute increase in prevalence than their female counterparts (10.1%, 95% CI: 9.9-10.2). The highest prevalence of HTN was estimated in black people according to both JNC7 (40.1%, 95% CI: 37.1-43.1) and ACC/AHA 2017 (54.7%, 95% CI: 51.6-57.7) guidelines, with an absolute difference of 14.6% (95% CI: 14.5-14.6) between guidelines. A similar highest prevalence of HTN was observed among people with a BMI of ≥30 according to both JNC7 (40.4%, 95% CI: 38.2-42.6) and ACC/AHA 2017 (57.7%, 95% CI: 54.4-61.0) guidelines, with an absolute difference of (16.3%, 95% CI: 16.2-18.4) between guidelines. The prevalence of HTN in people with diabetes was 80.4% (95% CI: 77.4%-83.4) based on ACC/AHA 2017 while it was 67.6% (95% CI: 65.1-69.9) according to JNC7 guideline with an absolute difference of 12.8% (95% CI: 12.3-13.5).

**Table 2:** Prevalence of hypertension among US adults in the NHANES 2017-18.

| **Characteristics** | **Hypertension by JNC 7**<br>**% (95% CI)** | **Hypertension by ACC/AHA 2017**<br>**% (95% CI)** | **Absolute**<br>**Difference**<br>**% (95% CI)** |
| --- | --- | --- | --- |
| **Overall** | 31.7 (28.7, 34.8) | 45.6 (43.0, 48.3) | 13.9 (13.5,14.3) |
| **Age (years)** |  |  |  |
| 18-39 | 7.0 (5.8-8.4) | 20.6 (18.7-22.7) | 13.6 (12.9-14.3) |
| 40-59 | 33.8 (31.3-36.3) | 51.2 (48.6-53.8) | 17.4 (16.7-17.5) |
| 60 and above | 63.0 (61.0-65.0) | 73.2 (71.1-75.3) | 10.2 (10.1-10.3) |
| **Gender** |  |  |  |
| Male | 33.1 (31.2-35.1) | 51.1 (49.0-53.2) | 18.0 (17.8-18.1) |
| Female | 30.3 (28.5-32.2) | 40.4 (38.4-42.4) | 10.1 (9.9-10.2) |
| **Race** |  |  |  |
| White | 32.7 (30.4-35.1) | 46.0 (43.5-48.4) | 13.3 (13.1-13.3) |
| Black | 40.1 (37.1-43.1) | 54.7 (51.6-57.7) | 14.6 (14.5-14.6) |
| Hispanic | 27.7 (23.5-32.1) | 38.5 (33.9-43.2) | 10.8 (10.4-11.1) |
| Other | 25.2 (23.1-27.4) | 41.7 (39.3-44.2) | 16.5 (16.2-16.8) |
| **BMI (kg/m<sup>2</sup>)** |  |  |  |
| <25 | 17.4 (15.3-19.6) | 29.1 (26.6-31.7) | 11.7 (11.3-12.1) |
| 25-29.99 | 33.0 (30.6-35.4) | 44.4 (41.8-47.0) | 11.4 (11.2-11.6) |
| >= 30 | 40.4 (38.2-42.6) | 57.7 (54.4-61.0) | 16.3 (16.2-18.4) |
| **Diabetic status** |  |  |  |
| No | 27.0 (25.6-28.4) | 41.1 (39.6-42.7) | 14.1 (14.0-14.3) |
| Yes | 67.6 (65.1-69.9) | 80.4 (77.4-83.4) | 12.8 (12.3-13.5) |

### 3.2 Hypertension control

Table 3 summarized the prevalence of CHTN and the absolute difference by age, sex, ethnicity, BMI, and diabetes status comparing the two guidelines. According to ACC/AHA 2017 guideline, the prevalence of CHTN was the highest among individuals aged 60 years and above (30.4%, 95% CI: 28.2-32.7), followed by 40-59 years (17.0%, 95% CI: 15.1-19.1), and 18-39 years (5.2%, 95% CI: 4.2-6.4). The corresponding prevalence based on JNC7 were 51.7% (95% CI: 49.3 −54.1) for 60 years and above age group, 47.0% (95% CI: 44.4-49.7) for 40-59 age group and 30.0% (95% CI: 27.7-32.3) for 18-39 years age group. The highest absolute difference between the two guidelines was 30.0% (95% CI: 29.3 −30.6) in the middle age group. According to the ACC/AHA 2017 classification 18.5% (95% CI: 16.9-20.1) male respondents had CHTN compared with 24.0% (95% CI: 22.3-25.7) of the female respondents. The prevalence of CHTN, according to JNC7, was 47.6% (95% CI: 45.5-49.6) among males and 48.8% (95% CI: 46.8-50.8) among females. Male (29.1%, 95% CI: 28.6 − 29.5) had a higher absolute increase in controlled prevalence than their female counterparts (24.8%, 95% CI: 24.5-25.1). The highest prevalence of CHTN was estimated in the white people according to both JNC7 (50.6%, 95% CI: 48.2-53.1) and ACC/AHA 2017 (22.8%, 95% CI: 20.8-24.9) guidelines, with an absolute difference of 27.8% (95% CI: 27.4-28.2) between guidelines. The highest rate of CHTN was observed among people with a BMI of ≥30 according to both JNC7 (53.2%, 95% CI: 50.1-55.4) and ACC/AHA 2017 (23.3%, 95% CI: 21.4-25.2) guidelines, with an absolute difference of 29.9%, 95% CI: 28.7-30.2) between guidelines. The prevalence of CHTN in people with diabetes was 60.8% (95% CI: 57.3-64.4) according to JNC7 guideline while was 32.9% (95% CI: 29.5-36.4) based on ACC/AHA 2017 guideline, with an absolute difference of 27.9% (95% CI: 27.8-28.0) between two guidelines.

**Table 3:** Control of hypertension among US adults in the NHANES 2017-18.

| **Characteristics** | **Hypertension control**<br>**by JNC 7**<br>**% (95% CI)** | **Hypertension control**<br>**by ACC/AHA 2017**<br>**% (95% CI)** | **Absolute Difference**<br>**% (95% CI)** |
| --- | --- | --- | --- |
| **Overall** | 48.2 (44.4, 52.0) | 21.0 (18.1, 24.2) | 27.2 (26.3, 27.8) |
| **Age** |  |  |  |
| 18-39 | 30.0 (27.7-32.3) | 5.2 (4.2-6.4) | 24.8 (23.5-25.9) |
| 40-59 | 47.0 (44.4-49.7) | 17.0 (15.1-19.1) | 30.0 (29.3-30.6) |
| 60 and above | 51.7 (49.3-54.1) | 30.4 (28.2-32.7) | 21.3 (21.1-21.4) |
| **Gender** |  |  |  |
| Male | 47.6 (45.5-49.6) | 18.5 (16.9-20.1) | 29.1 (28.6-29.5) |
| Female | 48.8 (46.8-50.8) | 24.0 (22.3-25.7) | 24.8 (24.5-25.1) |
| **Race** |  |  |  |
| White | 50.6 (48.2-53.1) | 22.8 (20.8-24.9) | 27.8 (27.4-28.2) |
| Black | 41.5 (38.6-44.5) | 17.5 (15.3-20.0) | 24.0 (23.3-24.5) |
| Hispanic | 40.7 (36.1-45.6) | 14.5 (11.3-18.2) | 26.2 (24.8-27.4) |
| Other race | 47.3 (44.8-49.8) | 19.3 (17.4-21.3) | 28.0 (27.4-28.5) |
| **BMI** |  |  |  |
| <25 | 33.7 (31.1-36.4) | 12.4 (10.7-14.4) | 21.3 (20.4-22.0) |
| 25-29.99 | 46.6 (44.1-49.2) | 22.1 (20.0-24.3) | 24.5 (24.1-24.9) |
| >= 30 | 53.2 (50.1-55.4) | 23.3 (21.4-25.2) | 29.9 (28.7-30.2) |
| **Diabetic status** |  |  |  |
| No | 44.1 (42.6-45.7) | 18.0 (16.8-19.2) | 26.1 (25.8-26.5) |
| Yes | 60.8 (57.3-64.4) | 32.9 (29.5-36.4) | 27.9 (27.8-28.0) |

## 4. Discussions

To our knowledge, this is one of the first studies comparing the prevalence of HTN and controlled HTN using JNC7 and ACC/AHA 2017 guidelines applied to NHANES 2017-2018 data. We have found that a third (31.7%) of the US population have HTN based on the older JNC7 guidelines. When the stricter 2017 ACC/AHA cut-offs were applied, this increases to almost half of the population (45.6%). Of the people who had HTN according to the JNC7, almost half (48.2%) had controlled blood pressure after commencing antihypertensive medication. Among those with HTN, according to ACC/AHA 2017 guidelines, only a fifth (21.0%) had controlled blood pressure levels after starting treatment. When blood pressure was assessed using both guidelines, the greatest absolute increase in rates of HTN and CHTN between JNC7 and ACC/AHA 2017 guidelines in 40-59 years old was 17.4% and 30.0%, respectively. These analyses suggest a higher prevalence of HTN and a lower level of controlled HTN when ACC/AHA 2017 guidelines are used. These findings have policy and funding implications for the management of HTN in the US.

We have found a high rate of HTN in the US population, irrespective of which guideline is used. Either a third (31.7%) or almost half (45.6%) of the US population have HTN based on the JNC7 or ACC/AHA 2017 guidelines, respectively. These are slightly higher than previous studies examining HTN in the US. For example, Guo et al., reported an overall age-adjusted prevalence of 29.5% (95% CI: 27.7 – 31.4) in 2009-2010, using the JNC7 cutoffs applied to NHANES 2009 – 2010 data [10]. This may reflect secular increases in HTN in the last decade associated with increases in risk factors such as obesity and the aging population [11–14]. However, these secular trends for increases in HTN may not be true in all age groups. In younger people (aged 20-39 years), the prevalence appears to have increased marginally between the period 2009-2010 and 2017-2018 (increase of 3.5%). In contrast, older adult’s HTN prevalence decreased by 3.7% between 2009-2010 to 2017-2018 [10]. This may reflect nuances in treatment approaches-older people are more likely to be screened, detected, and treated aggressively for HTN than younger people.

Our analysis has found discrepancies between HTN prevalence rates when the two guidelines are used. Differences in rates were found by age, sex, ethnicity, BMI, and diabetes status. The greatest discrepancy between the two guidelines was in the 40-59 years old age group. When JNC7 cutoffs were applied, a prevalence of HTN of 33.8% was found, whereas when ACC/AHA guidelines were used, the corresponding prevalence of HTN rose to 51.2 % (absolute increase=17.4%). This is higher than the discrepancy found in the reproductive-aged US women, where the absolute increase in rates of HTN between JNC7 and ACC/AHA 2017 was 8.7% [2]. The implication of our finding is that if the JNC7 guidelines were used, 17.4 people out of 100 aged 40-49 years would be classified as prehypertensive and, therefore, not receive treatment. Given the high burden of disease due to complications arising from untreated HTN, as well as the higher costs of untreated disease, these have very important public health implications.

These discrepancies between guidelines were even greater when the proportion of the population with CHTN was compared. According to ACC/AHA 2017 guideline, the prevalence of CHTN in 40-59 years old was 17.0%. The corresponding prevalence based on JNC7 was 47.0% (absolute difference between the two guidelines was 30.0%). Similar large discrepancies in rates of CHTN between guidelines between found for those with obesity (absolute difference = 29.9%), in males (29.1%), and those with diabetes (27.9%). These data suggest that far fewer patients have CHTN when assessed by ACC/AHA 2017 guidelines.

Our analysis confirms a very high prevalence of HTN among those with comorbidities, irrespective of the measure used. For example, the prevalence of HTN in people with diabetes was 80.4% based on ACC/AHA 2017 and 67.6% using JNC7 guidelines. This is comparable to other US studies in which prevalence rates of ∼70% in those with diabetes were found using JNC7 guidelines [2,4,15]. This cohort of patients are likely to benefit substantially from early drug treatment. For example, a systematic review and meta-analysis of blood pressure lowering in patients with type 2 diabetes found that BP lowering was associated with improved mortality and other clinical outcomes [16]. The greatest benefit was found in those with higher blood pressure at baseline. Our finding of a high prevalence of HTN in people with diabetes supports the use of medications for blood pressure lowering in these patients.

Our results are consistent with studies conducted in both high-income and low-middle-income countries that have applied the two guidelines. For example, a Canadian study examined the prevalence of HTN using the ACC/AHA 2017 guidelines and found that adoption of the ACC/AHA 2017 HTN guidelines would result in a near doubling in the prevalence of HTN in Canada [17]. Studies in the US, as well as in developing countries like Bangladesh and Nepal, found the ACC/AHA 2017 HTN guideline detected more participants as hypertensive when compared with the JNC7 guideline [2,18,19]. This suggests that the use of the ACC/AHA 2017 guidelines will have similar effects that of increasing the prevalence of HTN globally.

### 4.1 Strengths and Limitations

The strengths of the study are that it used a large, nationally representative dataset suggesting the findings have external validity. High-quality sampling also minimized sampling bias. Clinical variables were measured in mobile clinics by trained staff (e.g., blood pressure, weight, and height). Blood pressure measurements were made carefully using best practice techniques, including measurement of arm circumference to determine cuff size and the determination of maximum inflation level. The BP used in analyses was an average of three measures. A weighted analysis to adjust for the clustered sampling design of the NHANES survey was used to estimate the prevalence of HTN, which increased the precision of the findings. A limitation was that diabetes mellitus was self-reported. Also, we developed a secondary analysis from a publicly available survey, which limited the control of the variables, and stratifying for socioeconomic status was not undertaken in this analysis.

### 4.2 Public Health relevance and implications

HTN has a high burden of disease globally but is entirely preventable. Its surveillance and control must be a priority for the health systems around the world. Furthermore, new guidelines are important to continue with the screening and early detection of at-risk patients, such as those defined like pre-hypertension, to prevent complications across different populations. Guidelines with greater relevance to other groups, such as ethnic minorities, are also needed.

## 5. Conclusions

This analysis of contemporary, nationally representative NHANES data has shown that adoption of the ACC/AHA 2017 guidelines will result in an increase in the prevalence of HTN from a third to one half, compared with the JNC7 guidelines. Discrepancies exist between guidelines in the proportion of HTN and those with CHTN after starting antihypertensive treatment. The greatest discrepancies were in the middle-aged population (40-59 years old). While the application of stricter guidelines will result in a higher prevalence of those requiring treatment, it may result in cost savings by reducing complications arising from untreated HTN. Greater clarity is needed about which guidelines should be used to guide health policy in countries around the world.

## Data Availability

https://wwwn.cdc.gov/nchs/nhanes/Default.aspx

